# Coping skills, depression, and anxiety in medical students

**DOI:** 10.1101/2023.09.24.23295999

**Authors:** Sergio Baldassin, Mônica Levit Zilbermann, Tania Correa de Toledo Ferraz Alves, Anderson Sousa Martins-da-Silva, João Maurício Castaldelli-Maia, Luiz Antonio Nogueira-Martins

**Author notes:** Corresponding Author: Sergio Baldassin.

## Abstract

**Objective:** to describe coping strategies during medical education and their relationship with course periods, with gender, and depressive and anxiety range symptoms.

**Method:** the Ways of Coping Questionnaire (WCQ), the Beck Depression Inventory (BDI), and the Spielberger Trait Anxiety Inventory (STAI-T) were administered to 599 medical students from basic, intermediate and internship periods.

**Results:** The response rate was 80.3% (279 females and 190 males aged 21.9 ± 2.4 years). Belong to internship or basic/intermediate levels make a significant difference in choosing coping factors. Female students reported greater use of factors indicating a need for social support and fantasy. In contrast, male students reported greater use of factors such as substance use and self-control.

Twenty percent of students scored high on anxiety (STAI-T>49), whereas 7.6% scored on the depression range (>20) and 8.0% on the dysphoric range (16–20) of the BDI. Students at the normal range at the BDI and the STAI-T reported greater use of factors such as of social support and problem solving. The factor analysis of the WCQ showed that dysphoric, depressive, and high anxiety students reported greater use of factors indicating fantasy, search of relief and escape.

**Conclusion:** Dysphoria, depressive, and highly anxious medical students choose similar strategies to deal with stress related to medical graduation. Women and men exhibit distinct coping strategies related to medical education and training. Support and counseling services for medical students should take into consideration these depressive and anxiety ranges, and gender differences in coping strategies, with potentially important preventive and therapeutic implications.

## Introduction

Several studies have shown that medical students are particularly at risk for experiencing distress in association with the educational process itself.(1–3).(4) Stress-related problems among medical students are a matter of concern as they may impact medical training and future technical ability (5)

Systematic review of literature suggests that depression, anxiety, and burnout are frequent, (6) (7) however there were few studies exploring specially how and when the medical student choose among several coping strategies during their under-graduation program.

Curriculum contents,(8) hospital places to practice,(9) teachers’ attitudes(10, 11) and other factors(12) may increase anxiety and depressive symptoms thus affecting academic performance(13) and inducing dysfunctional coping(14–17) or other factors. The models (teachers and tutors)(18–20) the healthy handling of the difficulties(21, 22) with doctor-patients relationship and the perception of the own distress are important to learning and keep emotional healthy skills.(23–26)

General health,(27) sleep time,(28) quality of life, years of study at medical school(29) may increase distress and the present study aims to explore coping strategies during medical under graduation and their relationship to periods of course and depressive and anxiety symptoms.

There is evidence, also, suggesting that depressive and anxiety symptoms are significantly more frequent in females’ medical students as compared to males(7, 30–34). Particularly among first-year medical students, no initial differences in psychological adjustment between genders were noted, but by mid-year, female students had already developed more psychiatric symptoms, and were more dissatisfied with life.(31) It can be argued that stressors may affect women in specific ways, e.g., the female student may have difficulties reconciling the various expected roles (professional and personal) in addition to dealing with colleagues’ and instructors’ hostility in connection with their career choice as evidenced by remarks and jokes directed both towards them and female patients.

Nonetheless, there is evidence that female students are three times more likely to seek psychological support services and are also more able to recognize symptoms of stress than their male counterparts.(35)

Although it is acknowledged that gender might have an impact in the way medical students react to medical education(33, 36, 37) there is little research(38) exploring potential gender differences in coping strategies among medical students.

The present study aims to explore gender differences in coping during medical under graduation.

## Method

### Study Population

The target study population consisted of all 599 medical students (366 females; 61.1% and 233 males; 38.9%) regularly enrolled in a private Medical School..

The course was divided in three periods as usual in many countries(44): basic (1^st^ and 2^nd^ years), intermediate (3^rd^ and 4^th^ years) and internship (5^th^ and 6^th^ years).

### Procedure

The research protocol was approved by the institutional ethics committee(protocol number 026/2000). The research aims were explained to all regularly registered medical students present in the classrooms on the day of the survey (no prior announcement was made) who were then invited to participate. Those who agreed to participate signed the appropriate informed consent. Confidentiality was strictly ensured; the questionnaire was anonymous (only gender and age were indicated). No refusals occurred. In addition, subjects were provided with directions for accessing the psychological support service within the campus.

### Tools

#### Ways of Coping

A validate version of the Ways of Coping Questionnaire (WCQ) by Folkman and Lazarus (1985)(39) was used. The WCQ is self-administered 66-item measure, with responses ranging from 0 (=No) to 3 (=Extremely), that assesses attitudes and thoughts(40) to coping. For the present study, nine items were added to evaluate other attitudes commonly found in depressed and anxious subjects as follows: 67 – I used alcohol beverages, 68 – I used drugs,(41) 69 – I used tranquilizer,(42) 70 – I smoked cigarettes, 71 – I used vitamins; 72 – I used analgesics; 73 – I thought I would die; 74 – I tried to commit suicide,(43) and 75 – I thought I would withdraw from the medical course.

#### Depressive symptoms

Assessed using a validate version(45) of the Beck Depression Inventory (internal consistency in agreement with the literature: 0.81 for non-depressed subjects and 0.88 for depressed patients) with these cut-off points: normality (0–15), disphoric (16–20), depressive (>20).

#### Anxiety symptoms

Assessed using a validate version of Spielberger Trait Anxiety Inventory(45, 46) (STAI-T) with these cut-off points: mild anxiety (<33), moderate (33–49) and high (>49) were used too.

### Statistical analysis

Given the high number of items (75 with 4 alternatives each), a factor analysis of principal components using VARIMAX rotations of the Ways of Coping Questionnaire was conducted to yield fewer variables that could potentially explain the remaining factors.

To find possible relationship between multiple variables was used a classification tree method with minimal 100 cases before any division and not allowing <50 cases by groups.

A Student’s t-test was performed to compare factors resulting from the factor analysis between males and females. The non-parametric Mann-Whitney test was performed for factors that were outside the normal range.

A significance level of 5% was adopted for all tests.

## Results

The response rate was 80.3% (n = 481). Two hundred and seventy-nine female students (58.0%) and 190 male students (39.5%) completed the survey. Gender was not identified in 12 questionnaires (2.5%). The mean age was not significantly different between genders (22.0 ± 2.6 years for females and 21.9 ± 2.1 years for males; p= .303).

The absented 125 (19.7%) students were 50% women. 4.7% (83.3% male) of the first year, 29.7% of second year (73.7% male), 24.2% of third (67.7% female), 7 % of fourth (88.9% female), 21.9% of fifth (53.3% female), and 12.5% of sixth (56.3% female). The comparison between absentees and respondents regarding to gender (p=0.192) and age range < 20 years old and > 20 years old (p=0.281) were not statistically significant.

### Coping factors

The factor analysis of the WCQ produced 10 factors as follows:

**Factor 1 – Fantasy** (55-Wish that I can change what is happening or how I feel; 57-I daydream or imagine a better time or place than the one I am in; 58-Wish that the situation would go away or somehow be over with; 59-Have fantasies or wishes about how things might turn out.)
**Factor 2 – Problem solving** (26-I’m developing an action plan and following it; 49-I know what has to be done, so I am doubling my efforts to make things work; 46-I stand my ground and fight for what I want.)
**Factor 3 – Seeking social support** (8-Talk to someone to find out more about the situation; 42-Ask a relative or friend I respect for advice; 45-Talk to someone about how I’m feeling.)
**Factor 4 – Use of substances** (67-I drank alcoholic beverages; 68-I used drugs; 70-I smoked cigarettes.)
**Factor 5 – Comparison** (63-I thought about how a person I admire would handle this situation and used that as a model; 64-I tried to see things from the other persons’ point of view; 65-I reminded myself how much worse things could be.)
**Factor 6 – Search of relief** (22-I got professional help; 69-I used tranquilizers.)
**Factor 7 – Escape** (40-I avoided being with people in general; 47-Took it out on other people; 75-I thought of changing schools.)
**Factor 8 – Denial** (41-I didn’t let it get to me; refused to think too much about it; 44-Made light of the situation; refused to get too serious about it.)
**Factor 9 – Self-control** (54-I tried to keep my feelings from interfering with other things too much; 35-I tried not to act too hastily or follow my first hunch.)
**Factor 10 – Acceptance** (12-I went along with fate; sometimes I just have bad luck; 53-I accepted it, since nothing could be done.)

Throughout the course there is a difference in the use of the ten strategies of coping, problem solving was the most prevalent during basic, self control during intermediate and denial during internship.

Table 1 shows self-worth values and percentages of explained variance for these 10 factors. The final model was able to explain 61% of the total variance.

**Table 1:** Portion of variance explained by each factor identified in the factor analysis of the Ways of Coping Questionnaire.

| Items | Factors |  |  |  |  |  |  |  |  |  |
| --- | --- | --- | --- | --- | --- | --- | --- | --- | --- | --- |
|  | 1 | 2 | 3 | 4 | 5 | 6 | 7 | 8 | 9 | 10 |
| q57 | 0.83 | 0.02 | 0.09 | 0.00 | 0.09 | 0.04 | 0.05 | 0.03 | -0.09 | 0.06 |
| q58 | 0.81 | -0.01 | 0.07 | 0.01 | 0.00 | 0.08 | 0.17 | 0.09 | -0.02 | 0.00 |
| q59 | 0.80 | 0.09 | 0.03 | -0.01 | 0.10 | 0.05 | 0.01 | -0.04 | -0.01 | 0.13 |
| q55 | 0.70 | -0.11 | 0.08 | 0.03 | 0.09 | -0.07 | 0.20 | 0.04 | 0.14 | 0.04 |
| q26 | -0.06 | 0.79 | 0.10 | -0.01 | -0.03 | 0.00 | 0.00 | 0.00 | 0.05 | 0.08 |
| q46 | -0.04 | 0.72 | 0.13 | -0.03 | 0.07 | -0.08 | 0.02 | 0.01 | 0.09 | -0.12 |
| q49 | -0.02 | 0.72 | -0.01 | -0.06 | 0.17 | 0.03 | -0.03 | -0.14 | 0.00 | -0.03 |
| q39 | 0.19 | 0.59 | 0.03 | 0.06 | 0.14 | 0.14 | -0.20 | 0.23 | 0.08 | 0.01 |
| q45 | 0.03 | 0.03 | 0.86 | -0.06 | 0.05 | 0.03 | -0.04 | -0.09 | -0.01 | 0.05 |
| q42 | 0.10 | 0.03 | 0.82 | 0.00 | 0.15 | -0.05 | -0.10 | -0.05 | 0.07 | 0.07 |
| q8 | 0.14 | 0.20 | 0.69 | 0.00 | 0.03 | 0.08 | 0.09 | 0.02 | 0.01 | -0.09 |
| q68 | 0.05 | 0.03 | -0.07 | 0.77 | -0.02 | 0.11 | 0.04 | 0.10 | -0.08 | 0.06 |
| q67 | -0.03 | 0.00 | -0.06 | 0.77 | 0.09 | 0.07 | 0.10 | 0.10 | -0.08 | 0.12 |
| q70 | 0.00 | -0.08 | 0.06 | 0.66 | 0.06 | 0.02 | 0.10 | -0.07 | 0.09 | -0.11 |
| q65 | 0.06 | 0.02 | 0.04 | 0.11 | 0.73 | 0.02 | -0.02 | 0.11 | 0.01 | 0.08 |
| q63 | 0.13 | 0.19 | 0.06 | 0.02 | 0.72 | 0.06 | 0.04 | -0.08 | 0.03 | 0.01 |
| q64 | 0.10 | 0.12 | 0.17 | -0.01 | 0.65 | -0.06 | 0.13 | 0.07 | 0.28 | -0.16 |
| q22 | -0.13 | 0.01 | 0.17 | -0.14 | 0.16 | 0.74 | 0.08 | 0.08 | -0.10 | 0.06 |
| q69 | 0.13 | 0.00 | -0.02 | 0.13 | 0.11 | 0.72 | -0.02 | 0.08 | -0.20 | 0.08 |
| q74 | 0.03 | -0.04 | -0.09 | 0.22 | -0.15 | 0.48 | -0.01 | -0.11 | 0.16 | -0.02 |
| q21 | 0.16 | 0.09 | 0.03 | 0.10 | -0.17 | 0.46 | 0.19 | 0.05 | 0.18 | -0.27 |
| q75 | 0.08 | -0.11 | -0.01 | 0.09 | 0.14 | 0.03 | 0.69 | -0.11 | -0.04 | 0.21 |
| q47 | 0.23 | 0.11 | 0.10 | 0.21 | -0.06 | 0.00 | 0.65 | 0.11 | -0.19 | -0.02 |
| q40 | 0.25 | -0.16 | -0.21 | 0.04 | 0.06 | 0.17 | 0.60 | 0.23 | 0.09 | -0.05 |
| q41 | 0.03 | -0.04 | 0.07 | -0.05 | 0.06 | 0.06 | 0.14 | 0.80 | 0.04 | 0.15 |
| q44 | 0.07 | 0.02 | -0.20 | 0.18 | 0.01 | 0.01 | -0.03 | 0.72 | 0.08 | 0.04 |
| q35 | -0.03 | 0.08 | 0.06 | -0.12 | 0.15 | 0.04 | -0.02 | -0.07 | 0.72 | 0.14 |
| q54 | 0.01 | 0.13 | 0.01 | 0.06 | 0.07 | -0.10 | -0.13 | 0.26 | 0.68 | -0.05 |
| q12 | 0.15 | 0.01 | 0.07 | -0.01 | 0.04 | 0.02 | 0.02 | 0.19 | -0.06 | 0.80 |
| q53 | 0.12 | -0.09 | -0.06 | 0.11 | -0.09 | 0.00 | 0.22 | 0.02 | 0.36 | 0.67 |
| Self-worth | 2.8 | 2.2 | 2.1 | 1.9 | 1.8 | 1.6 | 1.6 | 1.5 | 1.4 | 1.4 |
| Variance | 9.3 | 7.4 | 7.1 | 6.2 | 5.9 | 5.4 | 5.3 | 5.1 | 4.7 | 4.6 |
| % Explained | 9.3 | 7.4 | 7.1 | 6.2 | 5.8 | 5.4 | 5.3 | 5.0 | 4.7 | 4.6 |
| % Accumulated | 9.3 | 16.7 | 23.8 | 30.0 | 35.9 | 41.3 | 46.6 | 51.6 | 56.3 | 61.0 |

Frequency of use of specific factors varied significantly by STAI-T and BDI scores (Tables 2 and 3). Problem solving and seeking of social support were the most reported coping strategies among students at normal BDI ranges, whereas for disphoric, depressive and high anxiety students the most common strategies were fantasy, search for relief, and escape.

**Table 2:**
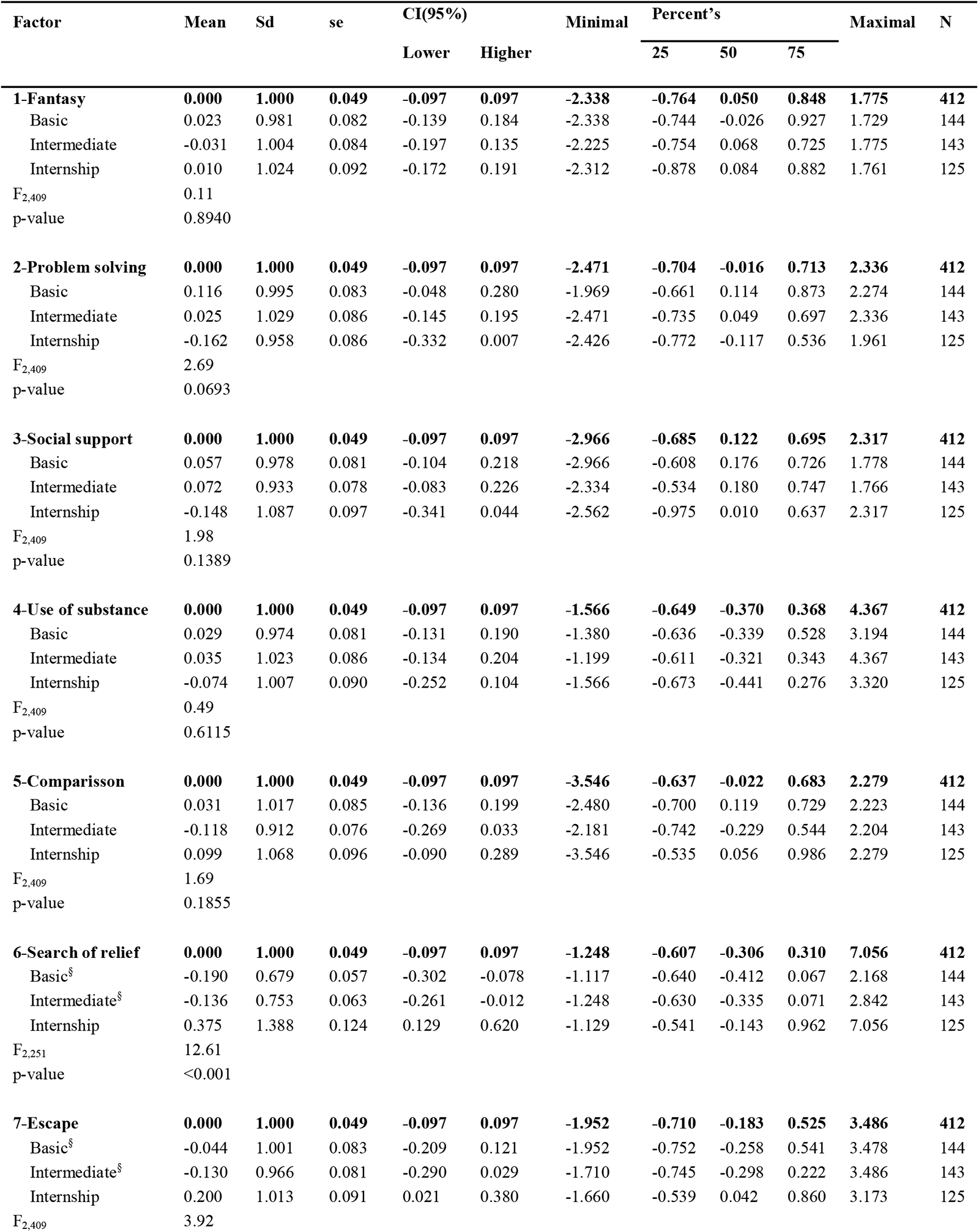
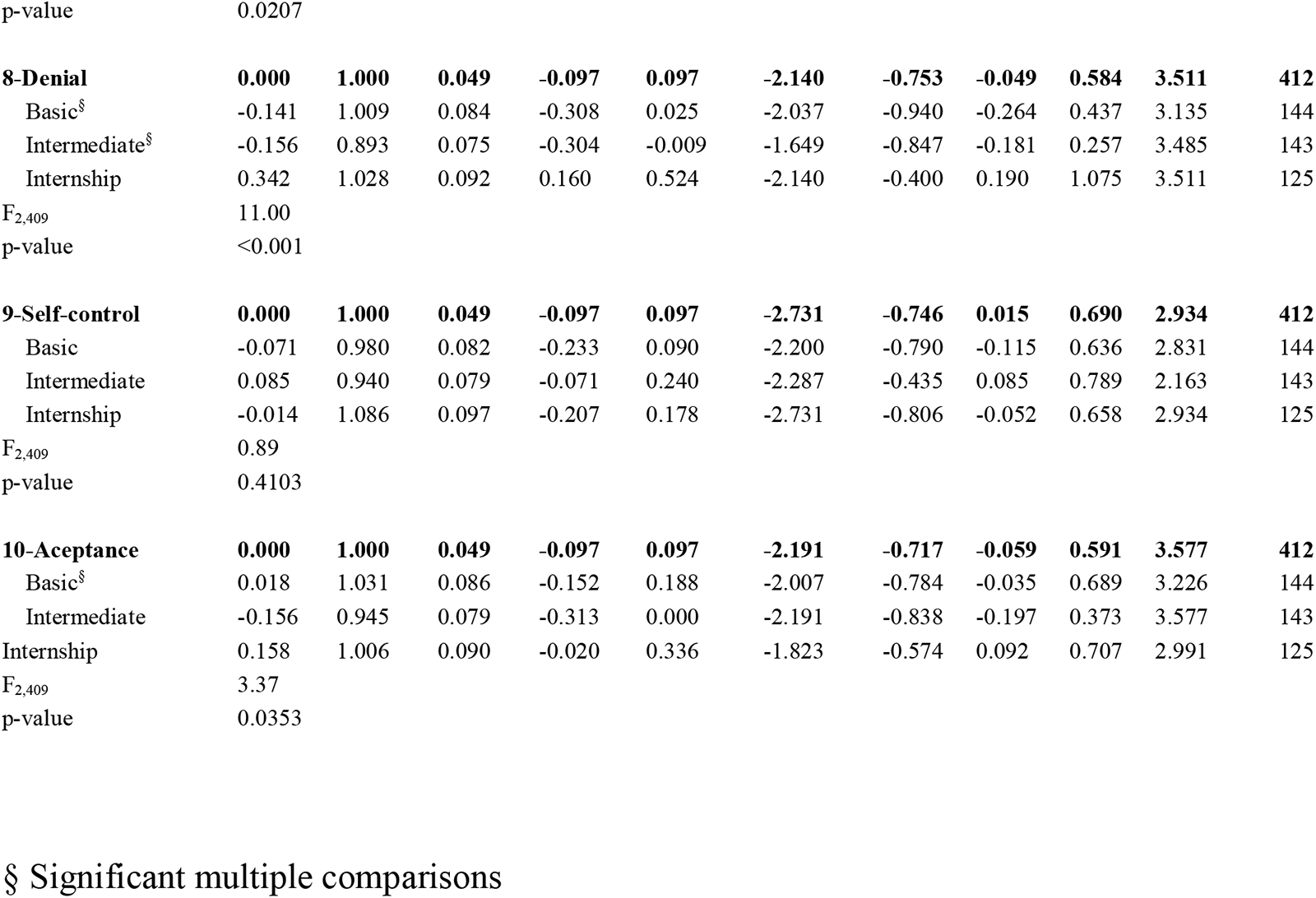
Factor analysis by coping factor and periods of course.

**Table 3:** Distribution by ranges of the Beck Depression Inventory and Spielberger Trait Anxiety Inventory of the means for 10 coping factors in medical students.

| Factor | STAI-T |  |  |  | BDI |  |  |  |  |
| --- | --- | --- | --- | --- | --- | --- | --- | --- | --- |
|  | Mild<br>33-49 | High<br>>49 | CI95% | P value | Normality<br><16 | Dysphoric<br>16-20 | Depressive<br>>20 | CI95% | P value |
| 1-Fantasy | -0.14 | 0.58 | -0.25:0.77 | <0.001 | -0.09 | 0.42 | 0.59 | -0.19:0.90 | <0.001 |
| 2-Problem solving. | 0.00 | 0.06 | -0.12:0.27 | 0.644 | 0.05 | -0.07 | -0.54 | -0.06:-0.22 | <0.001 |
| 3-Seek of social support | -0.04 | 0.07 | -0.15:0.33 | 0.448 | 0.07 | -0.31 | -0.65 | -0.03:-0.28 | <0.001 |
| 4-Use of substances* | -0.01 | -0.02 | -0.12:0.20 | 0.913 | -0.01 | 0.08 | 0.16 | -0.11:0.64 | 0.751 |
| 5-Comparisson | -0.05 | 0.18 | -0.16:0.42 | 0.070 | 0.01 | 0.17 | -0.14 | -0.09:0.31 | 0.482 |
| 6-Searck of relief* | -0.05 | 0.21 | -0.16:0.47 | 0.059 | -0.11 | 0.48 | 0.76 | -0.02:1.37 | 0.001 |
| 7-Escape* | -0.04 | 0.23 | -0.15:0.49 | 0.032 | -0.13 | 0.54 | 0.88 | -0.03:1.35 | <0.001 |
| 8-Denial* | 0.00 | 0.04 | -0.11:0.28 | 0.781 | -0.04 | 0.10 | 0.30 | -0.14:0.80 | 0.368 |
| 9 Self-control | 0.00 | 0.02 | -0.11:0.25 | 0.836 | 0.04 | -0.02 | -0.36 | -0.06:0.03 | 0.100 |
| 10-Acceptance | -0.05 | 0.05 | -0.16:0.30 | 0.422 | -0.02 | 0.27 | 0.23 | -0.13:0.64 | 0.146 |

t test for STAI-T and ANOVA for BDI.
Kruskall-Wallis (factors 4, 6, 7 and 8 for STAI-T and BDI).
Tukey and Dunett C (factors 1, 2, 3, 5, 9, and 10 for BDI).
\* Factors outside normal range after Kolmogorov-Smirnov test

Search of relief (p<0.001), escape (p=0.0207), denial (p<0.001), and acceptance (p=0.353) growing significantly from basic period to internship period (Table 2).

Table 4 shows self-worth values and percentages of explained variance for these 10 factors. The final model was able to explain 61% of the total variance.

**Table 4:** Portion of variance explained by each factor identified in the factor analysis of the Ways of Coping Questionnaire.

| Items | factors |  |  |  |  |  |  |  |  |  |
| --- | --- | --- | --- | --- | --- | --- | --- | --- | --- | --- |
|  | 1 | 2 | 3 | 4 | 5 | 6 | 7 | 8 | 9 | 10 |
| q57 | 0.83 | 0.02 | 0.09 | 0.00 | 0.09 | 0.04 | 0.05 | 0.03 | -0.09 | 0.06 |
| q58 | 0.81 | -0.01 | 0.07 | 0.01 | 0.00 | 0.08 | 0.17 | 0.09 | -0.02 | 0.00 |
| q59 | 0.80 | 0.09 | 0.03 | -0.01 | 0.10 | 0.05 | 0.01 | -0.04 | -0.01 | 0.13 |
| q55 | 0.70 | -0.11 | 0.08 | 0.03 | 0.09 | -0.07 | 0.20 | 0.04 | 0.14 | 0.04 |
| q26 | -0.06 | 0.79 | 0.10 | -0.01 | -0.03 | 0.00 | 0.00 | 0.00 | 0.05 | 0.08 |
| q46 | -0.04 | 0.72 | 0.13 | -0.03 | 0.07 | -0.08 | 0.02 | 0.01 | 0.09 | -0.12 |
| q49 | -0.02 | 0.72 | -0.01 | -0.06 | 0.17 | 0.03 | -0.03 | -0.14 | 0.00 | -0.03 |
| q39 | 0.19 | 0.59 | 0.03 | 0.06 | 0.14 | 0.14 | -0.20 | 0.23 | 0.08 | 0.01 |
| q45 | 0.03 | 0.03 | 0.86 | -0.06 | 0.05 | 0.03 | -0.04 | -0.09 | -0.01 | 0.05 |
| q42 | 0.10 | 0.03 | 0.82 | 0.00 | 0.15 | -0.05 | -0.10 | -0.05 | 0.07 | 0.07 |
| q8 | 0.14 | 0.20 | 0.69 | 0.00 | 0.03 | 0.08 | 0.09 | 0.02 | 0.01 | -0.09 |
| q68 | 0.05 | 0.03 | -0.07 | 0.77 | -0.02 | 0.11 | 0.04 | 0.10 | -0.08 | 0.06 |
| q67 | -0.03 | 0.00 | -0.06 | 0.77 | 0.09 | 0.07 | 0.10 | 0.10 | -0.08 | 0.12 |
| q70 | 0.00 | -0.08 | 0.06 | 0.66 | 0.06 | 0.02 | 0.10 | -0.07 | 0.09 | -0.11 |
| q65 | 0.06 | 0.02 | 0.04 | 0.11 | 0.73 | 0.02 | -0.02 | 0.11 | 0.01 | 0.08 |
| q63 | 0.13 | 0.19 | 0.06 | 0.02 | 0.72 | 0.06 | 0.04 | -0.08 | 0.03 | 0.01 |
| q64 | 0.10 | 0.12 | 0.17 | -0.01 | 0.65 | -0.06 | 0.13 | 0.07 | 0.28 | -0.16 |
| q22 | -0.13 | 0.01 | 0.17 | -0.14 | 0.16 | 0.74 | 0.08 | 0.08 | -0.10 | 0.06 |
| q69 | 0.13 | 0.00 | -0.02 | 0.13 | 0.11 | 0.72 | -0.02 | 0.08 | -0.20 | 0.08 |
| q74 | 0.03 | -0.04 | -0.09 | 0.22 | -0.15 | 0.48 | -0.01 | -0.11 | 0.16 | -0.02 |
| q21 | 0.16 | 0.09 | 0.03 | 0.10 | -0.17 | 0.46 | 0.19 | 0.05 | 0.18 | -0.27 |
| q75 | 0.08 | -0.11 | -0.01 | 0.09 | 0.14 | 0.03 | 0.69 | -0.11 | -0.04 | 0.21 |
| q47 | 0.23 | 0.11 | 0.10 | 0.21 | -0.06 | 0.00 | 0.65 | 0.11 | -0.19 | -0.02 |
| q40 | 0.25 | -0.16 | -0.21 | 0.04 | 0.06 | 0.17 | 0.60 | 0.23 | 0.09 | -0.05 |
| q41 | 0.03 | -0.04 | 0.07 | -0.05 | 0.06 | 0.06 | 0.14 | 0.80 | 0.04 | 0.15 |
| q44 | 0.07 | 0.02 | -0.20 | 0.18 | 0.01 | 0.01 | -0.03 | 0.72 | 0.08 | 0.04 |
| q35 | -0.03 | 0.08 | 0.06 | -0.12 | 0.15 | 0.04 | -0.02 | -0.07 | 0.72 | 0.14 |
| q54 | 0.01 | 0.13 | 0.01 | 0.06 | 0.07 | -0.10 | -0.13 | 0.26 | 0.68 | -0.05 |
| q12 | 0.15 | 0.01 | 0.07 | -0.01 | 0.04 | 0.02 | 0.02 | 0.19 | -0.06 | 0.80 |
| q53 | 0.12 | -0.09 | -0.06 | 0.11 | -0.09 | 0.00 | 0.22 | 0.02 | 0.36 | 0.67 |
| Self-worth | 2.8 | 2.2 | 2.1 | 1.9 | 1.8 | 1.6 | 1.6 | 1.5 | 1.4 | 1.4 |
| Variance | 9.3 | 7.4 | 7.1 | 6.2 | 5.9 | 5.4 | 5.3 | 5.1 | 4.7 | 4.6 |
| %Explained | 9.3 | 7.4 | 7.1 | 6.2 | 5.8 | 5.4 | 5.3 | 5.0 | 4.7 | 4.6 |
| %Explained | 9.3 | 16.7 | 23.8 | 30.0 | 35.9 | 41.3 | 46.6 | 51.6 | 56.3 | 61.0 |

Frequency of use of specific factors varied significantly by gender (Table 5). Fantasy and seeking of social support were the most reported coping strategies among female students, whereas for males the most common strategies were use of substances and attempts at self-control. Female and male students differed significantly regarding their use of fantasy, search of social support and use of substances (fantasy: p< .01; search of social support, use of substances: p< .001). The gender difference for the use of self-control strategies was non-significant (p= .149).

**Table 5:** Distribution by gender of the means for 10 coping factors in medical students.

| Factor | Gender |  |  |  |  |
| --- | --- | --- | --- | --- | --- |
|  | Female |  | Male |  | p value |
|  | mean | CI95% | mean | CI95% |  |
| 1-Fantasy | 0.12 | -0,01:0,24 | -0.19 | -0,34; -0,04 | 0.002 |
| 2- Problem solving | -0.02 | -0,14:0,10 | 0.04 | -0,12;0,20 | 0.549 |
| 3-Seeking social support | 0.21 | 0,10;0,33 | -0.34 | -0,50: -0,18 | <0.001 |
| 4-Use of substances * | -0.15 | -0,26: -0,04 | 0.24 | 0,07:0,42 | <0.001 |
| 5-Comparison | 0.02 | -0,10:0,14 | -0.03 | -0,18:0,13 | 0.658 |
| 6-Search of relief * | 0.02 | -0,09:0,14 | -0.04 | -0,21:0,13 | 0.374 |
| 7-Escape* | 0.06 | -0,06:0,19 | -0.08 | -0,24:0,07 | 0.082 |
| 8-Denial* | -0.01 | -0,14:0,11 | 0.04 | -0,12:0,20 | 0.808 |
| 9- Self-control | -0.05 | -0,18:0,07 | 0.09 | -0,07:0,25 | 0.149 |
| 10-Acceptance | -0.02 | -0,14;0,11 | 0.04 | -0,11;0,19 | 0.592 |

Student´s t-test (factors 1, 2, 3, 5, 9, 10 for Gender); Mann-Whitney test (factors 4, 6, 7, and 8 for Gender); \* - Factors outside normal range after Kolmogorov-Smirnov test

### Depression scores

Thirty-five subjects (7.6%) scored in the depression range in the BDI (mean score= 27.4 ± 8.6). Another 37 (8.0%) scored in the disphoric range (mean score= 17.8 ± 1.6). The remaining 388 (84.4%) subjects scored in the normal range (mean score= 6.6 ± 4.1). These groups showed significant differences (P<0.001).

### Anxiety scores

Eighty-nine (20.0%) subjects scored in the high anxiety range of the STAI (mean scored= 45.3 ± 4.7). Another 355 (79.8%) subjects scored in the moderate range (mean score= 43.6 ± 3.3) and only one subject (0.2%) scored in the low range with 32.0 points. These groups showed significant differences (P<0.001).

### Periods of Course

Using the classification trees, the method initially divides students into: node 1] basic/intermediate periods and node 2] internship period, who have distinct distributions of students (-2obs =20.59; p-value < 0.001).

For students of the basic/intermediate periods (node 1), the **factor 7 scape** were the most important variable (-_2obs_=25.00; p-value = 0.006) generating three groups respectively with **factor values 7 scape:** node 3] less than or equal to 0.10462, node 4] between 0.10642 and 1.37459, and node 5] above 1.37459, constituted by 89.5% of the students in the normal BDI range, 5.7% in the dysphoria range and 4.8% in the depression range.

For the students of the internship period (node 2), ways of coping **factor 9 self-control** were the most important variable (-_2obs_=12.08; p-value = 0.045) generating two groups, respectively: node 6] with values of **factor 9 self-control** less than or equal to 0.52682, and node 7] above 0.52682, constituted by 73.1% in the normal **BDI** range, 13.1% in the dysphoria range and 13.8% in the depression range.

Thus, the decision tree indicated the formation of five distinct groups of students regarding **BDI**. The student’s course period was the most important variable to discriminate students according to the **BDI**, and the students at the internship were more dysphoric and depressed.

The students of the internship with values, in the **factor 9 self-control**, above 0.52682 (node 7) presented the highest percentage of dysphoric (23.3%) and 6.7% in the depression range. The group with values lower than or equal to 0.52682 (node 6) presented 5.9% in the dysphoria range, and 18.8% of students in the depression range.

The group of the basic/intermediate periods with values in the **factor 7 scape** above 1.37459 (node 5) presented 13.8% of students in the depression range. The group with values of **factor 7 scape** between 0.10642 and 1.37459 (node 4) presented the second highest percentage of students in the normal range for **BDI** (86.2%).

Finally, the group of students from the basic/intermediate periods with values in coping **factor 7 scape** lower than or equal 0.10642 (node 3) presented the highest percentage of students in the normal range (96.0%) of the **BDI** and the lowest percentages of dysphoria (2.3%) and depression (1.7%).

For **STAI-T** ranges (Figure 2) the classification tree showed significant association (^2^obs =25.352; df=2; p value< 0.001) only for coping **factor 1 fantasy** generating two groups: node 1: scores ≤-0.313, node 2: scores >-0.3

**Figure 1.**
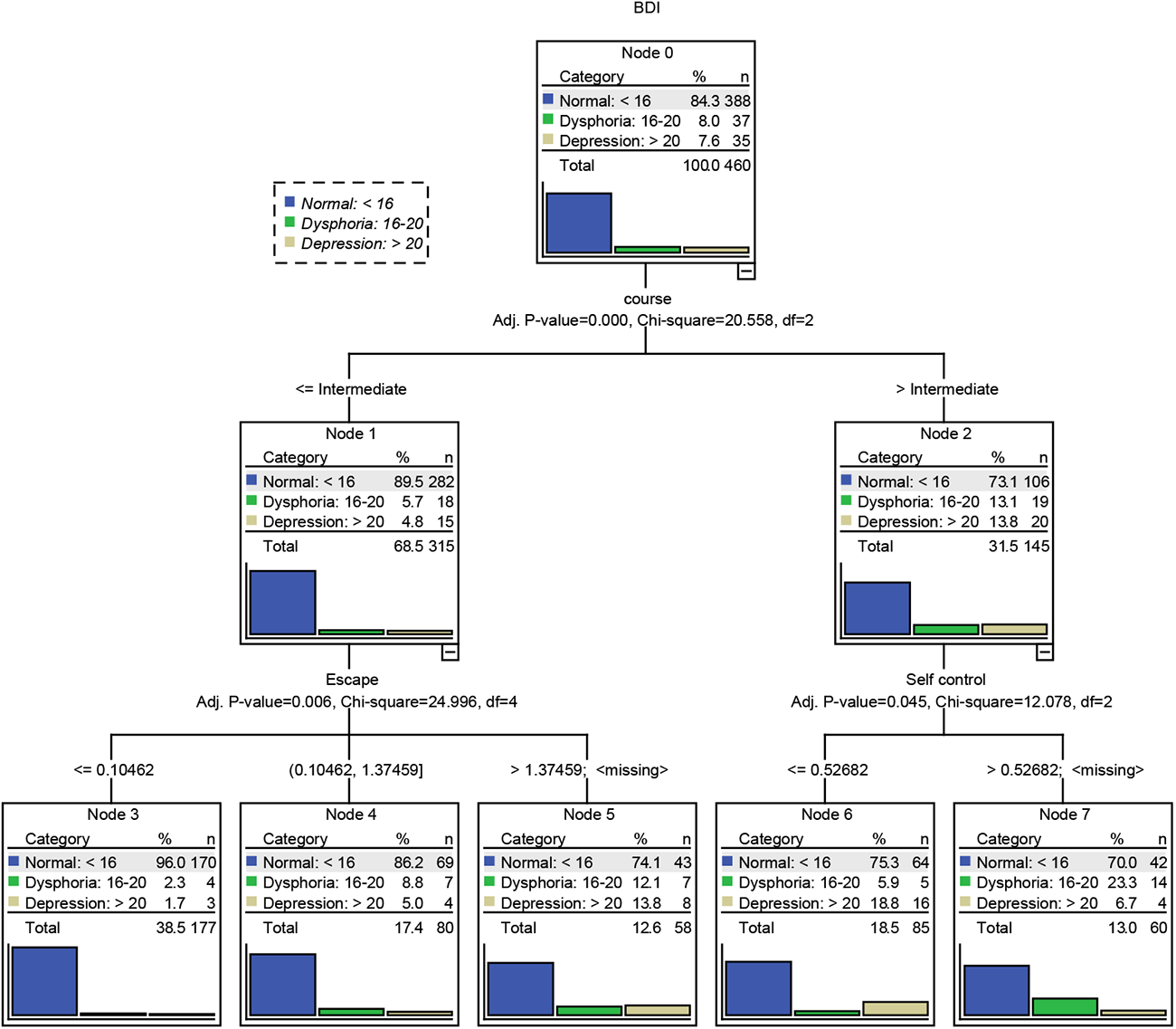
associated factors for BDI ranges among medical students

**Figure 2.**
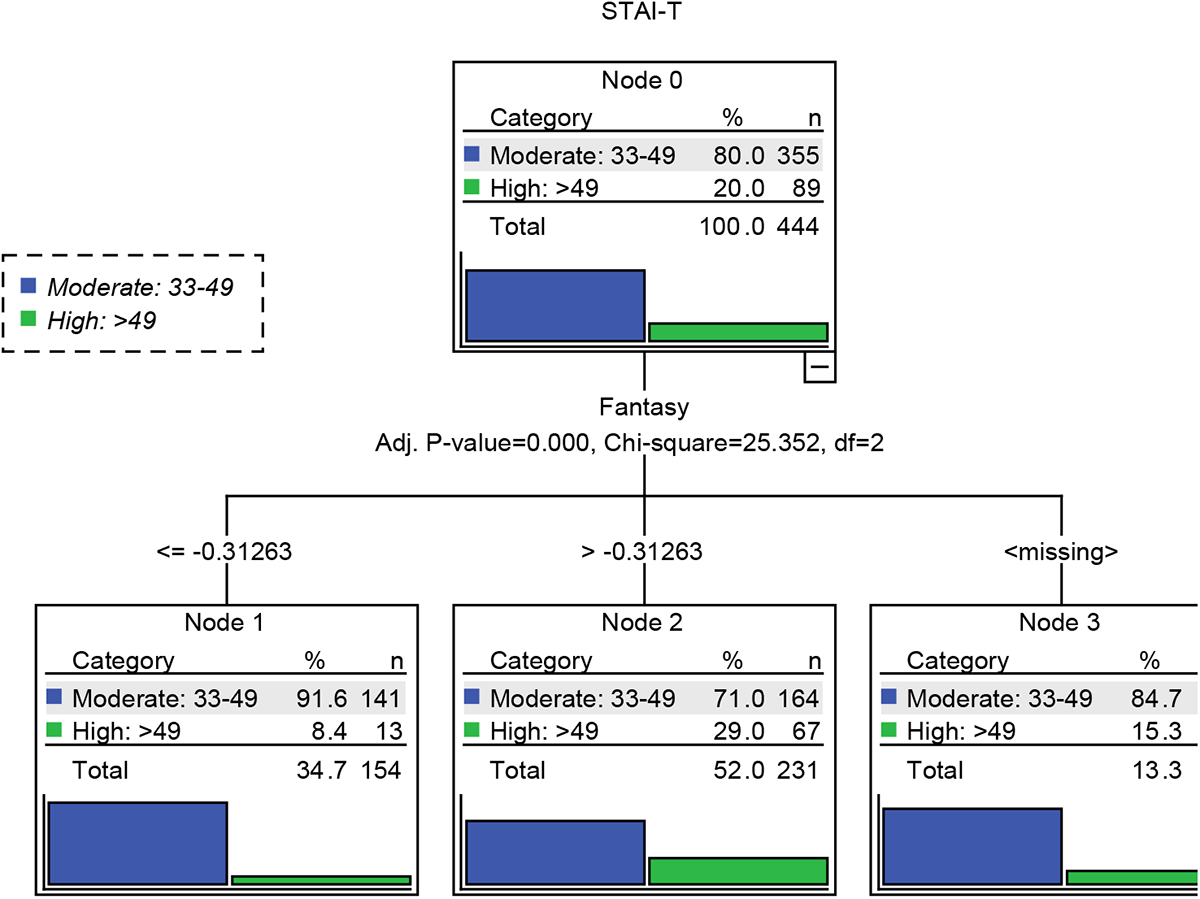
associated factors for Spielberger Traits Anxiety Inventory among medical students.

## Discussion

The response rate obtained in this survey can be considered high in comparison with other studies with medical students.(6) Another strength of this survey was the ability to evaluate students in all six years of medical graduation.

Limitations of this study include the loss of data from absent students, anonymous and self-application questionnaires, and a small sample. It is possible that absence from classroom itself is a way of coping.

The factor analysis of the WCQ yielded 10 distinct factors similarly to those obtained by Savóia et al.(39) Some of these factors encompass strategies that are probably more functional (e.g. problem-solving, search of social support, comparison) than others that could be considered somewhat dysfunctional (e.g. fantasy, use of substances, escape, denial), while some of them may be doubtfully functional (e.g. search of relief, attempts at self-control, acceptance).

In this sense, our results suggest that in this sample of students at the normal range for BDI and low and moderate range for STAI-T the most frequent coping strategies are a combination of functional factors (solving problems and search of social support), whereas dysphoria, depressive, and high anxiety students used more dysfunctional (fantasy and escape) and doubtfully functional (search of relief) strategies. From the basic/intermediate periods to the internship period, there was a three times increase in depressive symptoms and 2.5 times for dysphoria.

Regarding gender, the most frequent coping strategy is a combination of a functional factor (search of social support) with a dysfunctional one (fantasy) for female students, while male students’ strategy combines both a dysfunctional factor (use of substances) and one of doubtful functionality (self-control).

The fact that female students make more use “search of social support” may indicate their being more likely to seek psychiatric and psychological consultation than male students, as reported by other authors.(3, 47) On the other hand, the use of substances is a major coping strategy by male students(48, 49) being of particular concern, since this may mask emotional distress, thereby delaying appropriate assessment and treatment.

## Conclusion

These results indicate that emotional state (depressive and anxiety symptoms), gender, and periods of course need to be considered in developing preventive and support techniques (educators, teachers, psychologists, counselors, social workers, and pedagogues) to stimulate healthy coping among medical students. Future research should explore the development of specific approaches to enhance the balanced use of functional coping strategies.

## Data Availability

All data produced in the present study are available upon reasonable request to the authors.

## Acknowledgement

We are grateful to the comments carried out by prof. Jair Mari (Federal University of Sao Paulo).

